# Identification of actionable genetic variants in 4,198 Scottish volunteers from the Viking Genes research cohort and implementation of return of results

**DOI:** 10.1101/2024.11.01.24316571

**Authors:** Shona M. Kerr, Lucija Klaric, Marisa D. Muckian, Kiera Johnston, Camilla Drake, Mihail Halachev, Emma Cowan, Lesley Snadden, John Dean, Sean L. Zheng, Prisca K. Thami, James S. Ware, Gannie Tzoneva, Alan R. Shuldiner, Zosia Miedzybrodzka, James F. Wilson

## Abstract

The benefits of returning clinically actionable genetic results to participants in research cohorts are accruing, yet such a genome-first approach is challenging. Here, we describe the return of such results in two founder populations from Scotland. Between 2005 and 2015, we recruited >4,000 adults with grandparents from Orkney and Shetland into the Viking Genes research cohort. Return of genetic data was not offered at baseline, but in 2023 we sent invitations for consent to return of actionable genetic findings to participants. We generated exome sequence data from 4,198 participants, and used the ACMG v3.2 list of 81 genes, ClinVar review and pathogenicity status, plus manual curation, to develop a pipeline to identify potentially actionable variants. We identified 104 individuals (2.5%) carrying 108 actionable genotypes at 39 variants in 23 genes, and validated these. Working with the NHS clinical genetics service, which provided genetic counselling and clinical verification of the research results, and after expert clinical review, we notified 64 consenting participants (or their next of kin) of their actionable genotypes. Ten actionable variants across seven genes (*BRCA1, BRCA2, ATP7B, TTN, KCNH2, MUTYH, GAA* ) have risen 50 to >3,000-fold in frequency through genetic drift in ancestral island localities. Viking Genes is one of the first UK research cohorts to return actionable findings, providing an ethical and logistical exemplar of return of results. The genetic structure in the Northern Isles of Scotland, with multiple founder effects, provides a unique opportunity for a tailored approach to primary and secondary prevention through genetic screening.

## Introduction

Where exome or genome sequencing is clinically indicated, the opportunity arises to report additional incidental findings with the aim of diagnosis and pre-emptive clinical intervention. The American College of Medical Genetics and Genomics (ACMG) has led this approach and annually updates a benchmark list of genes for report^1^. The list currently has 81 genes, which meet strict criteria: there must be strong evidence that certain variants within the gene cause disease, and there must be effective medical intervention (for example screening or medication) which can be offered to the carrier. The great majority of the genes are associated with either inherited cancer predisposition, cardiovascular risk or metabolic diseases. These conditions are mostly inherited in an autosomal dominant fashion, but variants in genes causing autosomal recessive disease which are only actionable if homozygous or compound heterozygous are also present in the ACMG v3.2 list. Although there has not yet been universal international adoption of this approach, the ACMG list is the most widely used.

The consent obtained from participants in many research cohorts to date, including UK Biobank (UKB)^2^ , does not allow the return of results about actionable variants (AVs) or genotypes. However, some newer US cohorts, such as *All of Us,* are committed to making their research results, including hereditary disease risk, accessible to participants^3^. Geisinger’s MyCode Community Health Initiative provides a model for “genome-first” care^4^, and an international policy on return of results acknowledges the potential medical benefits to the individuals who are participating in genomics research^5^.

The populations of the Northern Isles of Scotland – the Orkney and Shetland archipelagos - are the most genetically isolated of all British and Irish populations, with the highest degree Norse admixture, and an enrichment of rare and low frequency functional variants^6–9^. Viking Genes (<u>viking.ed.ac.uk</u>) comprises a set of Northern and Western Isles population cohort studies, aiming to explore genetic causes of disease – the Orkney Complex Disease Study (ORCADES), and VIKING I, II and III. ORCADES^9^ and VIKING I^10^ together contain a rich data resource of more than 4,000 deeply phenotyped and exome sequenced research subjects, nearly all with three or four grandparents from Orkney, or Shetland, respectively. In total, Viking Genes has over 10,000 participants, all of whom consented to analysis of their DNA, and to be contacted at a later date in connection with future ethically approved studies. We have previously reported founder variants in *KCNH2*, *BRCA1* and *BRCA2* in Shetland and Orkney populations^10–12^, with variants tracing back to individual island ancestries.

In Viking Genes, all cohort members were offered the option of consent to return of actionable genetic results. Building on a method developed by Kelly *et al*^13^ to screen 130,048 adults in the Geisinger MyCode project, we used the ACMG v3.2 list^1^ and variant classification information from the public database ClinVar^14^ to code automated filters to identify potential actionable variants within 4,198 members of the Viking Genes cohort. Here, we describe the analysis pipeline, summarize actionable results, the process of returning these, and some challenges associated with implementation of these procedures at scale. We provide evidence of multiple drifted actionable founder variants that could form the basis for a future targeted Northern Isles population genetic screening programme.

## Subjects and Methods

### Research Volunteer Recruitment and DNA Collection

Recruitment to ORCADES took place from 2005-2011^11^, and to Viking I (Viking Health Study – Shetland) from 2013-2015^10^. Blood or occasionally saliva samples from participants were collected, processed and stored using standard operating procedures and managed through a laboratory information management system at the Edinburgh Clinical Research Facility, University of Edinburgh. Recruitment to VIKING II and VIKING III took place online from January 2020 to March 2023^15^.

### Viking Genes Pedigree Information

Records of the births, marriages and deaths in Orkney and Shetland are held at the General Register Office for Scotland (New Register House, Edinburgh). These records, along with relationship information obtained from study participants, Orkney and Shetland family history societies, and genealogies available online, were used to construct pedigrees of study participants using RootsMagic software (S&N Genealogy Supplies), which was then amended to reflect the genetic kinship between individuals using genotype data.

### Genotyping

DNA from all participants was used for genome-wide genotyping on the GSA BeadChip (Illumina) at the Regeneron Genetics Center. Monomorphic genotypes and genotypes with more than 2% of missingness and Hardy-Weinberg equilibrium (HWE) p<10^-6^ were removed, as well as individuals with more than 3% of missingness.

### Exome Sequencing

Quality controlled exome sequence data sets were prepared at the Regeneron Genetics Center, following the process detailed for UKB^2^, with 94.6% of targeted bases having >20X coverage.

### Pipeline Development

To find carriers of actionable variants, we first extracted all variants that map to genes in the ACMG v3.2 list from the whole exome sequences (WES)^1^. These variants were annotated with the dbsnp version 151 using the bcftools 1.9 annotate function, using CHROM,POS,ID,REF,ALT columns. The dbsnp 151 data was downloaded from https://ftp.ncbi.nlm.nih.gov/snp/organisms/human_9606_b151_GRCh38p7/VCF/ (00-All.vcf.gz file) in March 2022. To assess their pathogenicity, these variants were then merged with the ClinVar data. The variant_summary.txt.gz file of the Clinvar data was downloaded on 18^th^ March 2024, from https://ftp.ncbi.nlm.nih.gov/pub/clinvar/tab_delimited/. Merging by chromosome, position (genome build GRCh38), reference (REF) and alternate (ALT) alleles was necessary, since rsids can be multiallelic - containing multiple variants at the same genomic position, including different nucleotide substitutions, for example tri-allelic SNPs (A>C, A>G) as well as indels, and therefore merging solely by rsids might result in merging different variants.

Some of the variants present in the exomes were not identical to mutations listed in ClinVar, but result in the same amino-acid change as variants in ClinVar and thus the same consequence, commonly a frameshift with comparable functional effects. Therefore, we also merged the variants from the WES with ClinVar data based on the change of the amino-acid that the variant causes, instead of the CHR, POS, REF, ALT. Furthermore, if the following hypothetical indel occurred, TCATCTA > TCATCTCTA, it could be named either as c.4insCT or c.5insTC or c.6insCT, causing problems for merging the data using either of the two approaches. This is illustrated by the real variant rs886044536, *TTN* c.93396_93400del AGCTT ([MIM: 188840], Results). This is a deletion: GCCAAGCTAAGACT > GCCAAGACT which can be rendered either as GCCAAG[CTAAG/*]ACT or as GCC[AAGCT/*]AAGACT.

After merging, we next filtered the output using the ClinVar annotations to retain only pathogenic or likely pathogenic variants (P/LP), that had status 1 star or higher, that is with assertion of pathogenicity. Moreover, the variant class had to match the disease-causing variant classes for that gene, e.g. only loss-of-function (truncating) variants for titin (*TTN*). Variants were also subject to manual review before listing as potentially actionable.

### Whole Genome Sequencing

Whole genome sequencing (WGS) data is available for 1,300 members of the ORCADES cohort^16^ and 500 members of the Viking I cohort^17^. These datasets were used for look-ups, as part of the verification process of the actionable variant results obtained from exome sequencing.

### Sanger Sequencing

Pre-designed primer pairs for PCR and Sanger sequencing were selected from a collection that covers over 95% of human coding exons (Primer Designer, ThermoFisher Scientific). If validated primers were not available, primer pairs were designed using Primer3 software (ThermoFisher Scientific) and checked using UCSC In Silico PCR^18^. A standard protocol for PCR and Sanger sequencing on an Applied Biosystems 3730xl Genetic Analyser was applied across all samples. Output files were analysed using Sequencher DNA sequence analysis software version 5.4.6 (Gene Codes Corporation).

Validation of heterozygous short insertions/deletions (indels) can be challenging with Sanger sequencing. Although still considered the gold standard, the sequencing ladders derived from, for example, the deleted and non-deleted alleles become out of synchronicity with one another (by the length of the indel in base-pairs), leading to double sequence after the indel in each direction. Illumina short read sequencing, as used to generate the exome data, does not suffer from this problem. A further issue is that certain indels with short repeats within or near them (or in low complexity DNA) can be rendered in multiple ways in cDNA nomenclature, e.g. *TTN* c.93396_93400delAGCTT (see above). More common are single base indels altering the length of homopolymers, which can be annotated as a change at any position in, for example, the polyA tract. Chromosome positions are affected in the same way, and so when interrogating ClinVar, we performed a secondary merge using the protein sequence nomenclature, to pick up such instances.

### Clinical review of potentially actionable genotypes

All potentially actionable variants and genotypes presented in Table S1 other than those noted as not determined were discussed with the NHS clinical team in order to agree suitability for return of results to participants. Local clinical knowledge in NHS Grampian was used as part of this review process. *Electronic Health Record (EHR) Data Linkage*

This work used data provided by patients and collected by the NHS as part of their care and support, reused with permission from Public Health Scotland. NHS routine datasets linked to ORCADES and VIKING I participants in July 2021, including SMR01, the general/acute hospital inpatient and day case dataset of episode level data, and SMR00, outpatient appointments and attendances, were accessed using a secure process, as described previously^11^.

## Results

### Exome sequencing and validation

Exome sequences were generated from 4,198 participants from the Viking Genes cohort using industry-standard protocols as described in the Methods section. The exome sequence calls are of high quality, but it is important to verify them in the research laboratory, before considering the process of notifying a potential carrier. We therefore attempted to validate 149 potential actionable variant genotypes: 77 by Sanger sequencing, 48 by WGS and 24 by both sequence analysis methods. The total of 149 includes a number of heterozygous carriers of recessive variants in genes on the ACMGv3.2 list. 147 out of the 149 assessed were validated (98.7%). Notably, both of the variants that failed (one by Sanger sequencing alone, and one by both Sanger sequencing and WGS) were only 2 kb apart in the same gene, *MSH2* [MIM: 609309]. Manual review of the exome read stacks suggests that these variants are both false positives, with low read numbers and increased noise: they were not included in the total counts of actionable variants found.

One variant, in *DES* [MIM: 125660], c.973C>T, was found to be most likely a somatic mosaic in the sample of venous blood and so was not included in our counts and not returned. In Sanger sequencing from forward and reverse strands, only a very weak T was observed, and the read counts in WGS were 19 C and 1T. In 85X exome data, the total read counts (sum of forward and reverse) are 72C, 13T, for a variant allele distribution of 15%.

### Pipeline for identification of potential actionable variants

Exome sequence data from 2,090 ORCADES participants (820 male and 1,270 female) and 2,108 VIKING I participants (843 male and 1,265 female) passed all sequence and genotype quality control thresholds (Methods). The data were run through our pipeline (Methods), a key component of which is the ClinVar resource (www.ncbi.nlm.nih.gov/clinvar/). ClinVar aggregates information about genomic variation and its relationship to human health and allocates a clinical significance category to each variant^14^.

In total, 59 potential actionable variants were observed, consisting of 27 singletons (Supplemental Table S1) and 32 non-singleton variants (Tables 1 and 2). Half of the singleton variants (13/27) were observed in participants with one non-islander parent, and a further 4/27 were in volunteers with one non-islander grandparent, for a total of 63% of singletons with at least one quarter non-Northern Isles DNA. The remaining 10 singletons presumably arose by mutation in the last few generations and are thus very rare, or were brought into the islands by non-parental events. The variants were found in 26 different genes. For three of these genes, *BTD* [MIM: 609019], *CASQ2* [MIM: 114251] and *RPE65* [MIM: 180069], each carrier only had a single copy of a variant that acts recessively, thus not an actionable genotype. In this paper we are careful to distinguish between actionable variants (from the ACMG list) and actionable genotypes, the combinations of such variants which are actionable, for example compound heterozygotes or homozygotes for autosomal recessive conditions. Altogether, using the 81-gene ACMG 3.2 list, we observed actionable genotypes for 104 individuals (2.5% of the population) carrying 108 actionable genotypes at 39 of the variants, across 23 genes. No X-linked actionable variants were found.

**Table 1.**
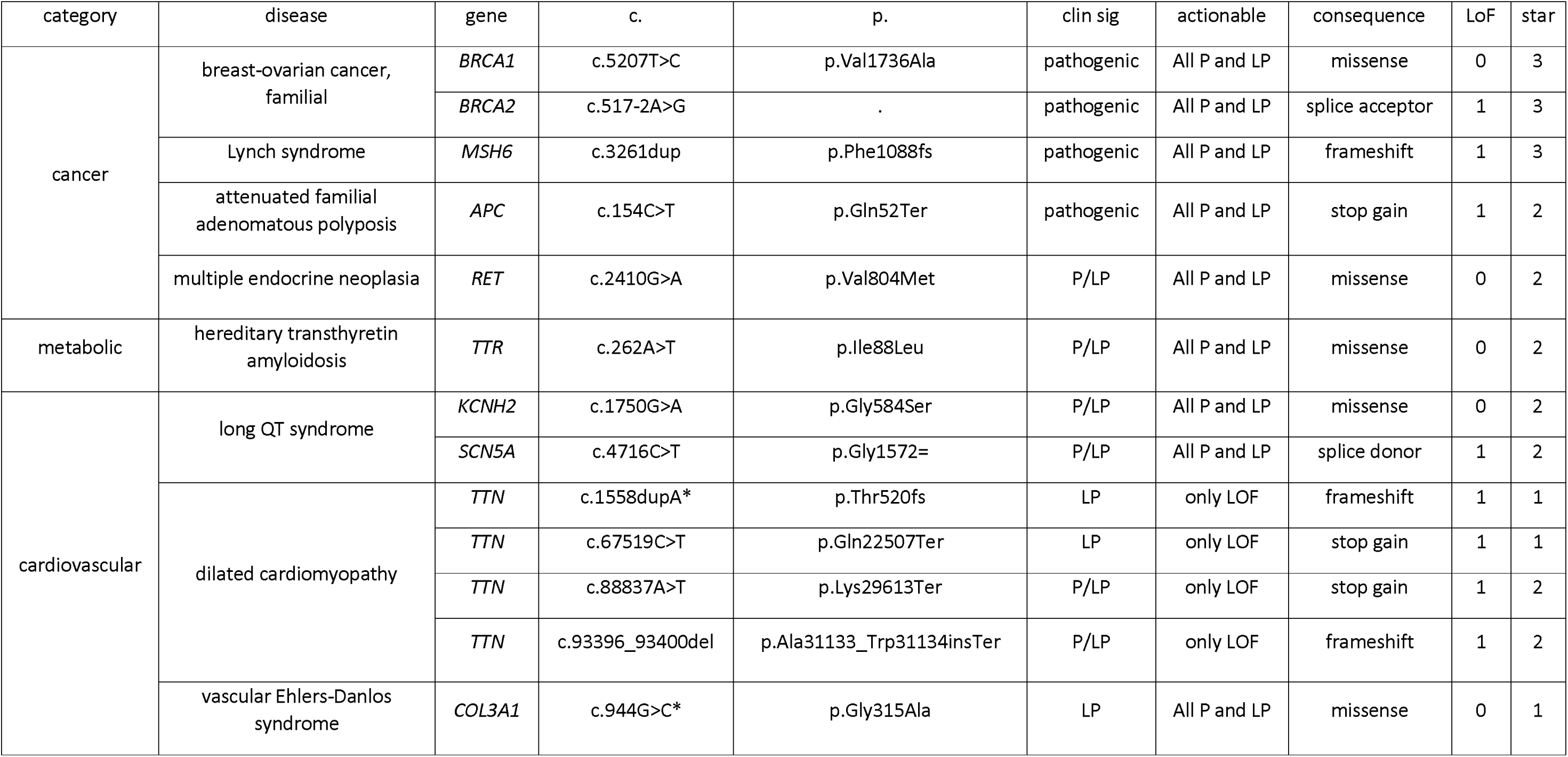
Summary of non-singleton autosomal dominant actionable variants in the cohort of 4,198 participants. The diseases are grouped into the categories of cancer, metabolic and cardiovascular. c., coding sequence change; p., protein sequence change; clin sig, clinical significance accordin g to ClinVar; LP, likely pathogenic; P/LP, pathogenic/likely pathogenic; actionable, variant categories considered to be actionable; LOF, loss of function; consequence, the molecular consequence of the variant; star, ClinVar star status. TTN c.1558dupA does not appear in ClinVar, so the star status and clinical significance are given fo r a variant causing the identical amino-acid change. *These variants were not considered actionable by the NHS Clinical Genetics team and were not returned.

| category | disease | gene | c. | p. | clin sig | actionable | consequence | LoF | star |
| --- | --- | --- | --- | --- | --- | --- | --- | --- | --- |
| cancer | breast-ovarian cancer, familial | *BRCA1* | c.5207T>C | p.Val1736Ala | pathogenic | All P and LP | missense | 0 | 3 |
|  |  | *BRCA2* | c.517-2A>G | . | pathogenic | All P and LP | splice acceptor | 1 | 3 |
|  | Lynch syndrome | *MSH6* | c.3261dup | p.Phe1088fs | pathogenic | All P and LP | frameshift | 1 | 3 |
|  | attenuated familial adenomatous polyposis | *APC* | c.154C>T | p.Gln52Ter | pathogenic | All P and LP | stop gain | 1 | 2 |
|  | multiple endocrine neoplasia | *RET* | c.2410G>A | p.Val804Met | P/LP | All P and LP | missense | 0 | 2 |
| metabolic | hereditary transthyretin amyloidosis | *TTR* | c.262A>T | p.Ile88Leu | P/LP | All P and LP | missense | 0 | 2 |
| cardiovascular | long QT syndrome | *KCNH2* | c.1750G>A | p.Gly584Ser | P/LP | All P and LP | missense | 0 | 2 |
|  |  | *SCN5A* | c.4716C>T | p.Gly1572= | P/LP | All P and LP | splice donor | 1 | 2 |
|  | dilated cardiomyopathy | *TTN* | c.1558dupA* | p.Thr520fs | LP | only LOF | frameshift | 1 | 1 |
|  |  | *TTN* | c.67519C>T | p.Gln22507Ter | LP | only LOF | stop gain | 1 | 1 |
|  |  | *TTN* | c.88837A>T | p.Lys29613Ter | P/LP | only LOF | stop gain | 1 | 2 |
|  |  | *TTN* | c.93396_93400del | p.Ala31133_Trp31134insTer | P/LP | only LOF | frameshift | 1 | 2 |
|  | vascular Ehlers-Danlos syndrome | *COL3A1* | c.944G>C* | p.Gly315Ala | LP | All P and LP | missense | 0 | 1 |

**Table 2.**
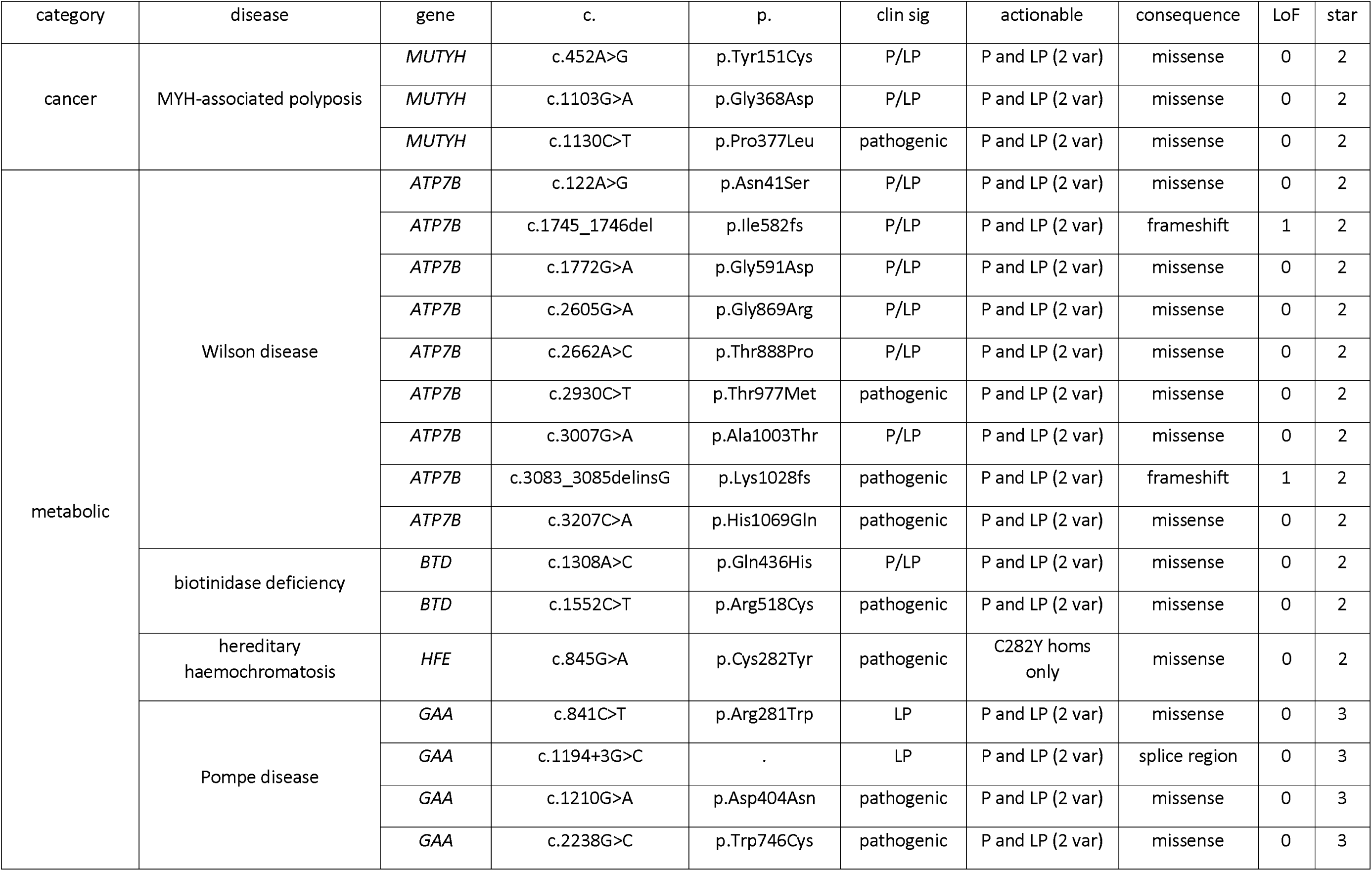
Summary of non-singleton autosomal recessive actionable variants in the cohort of 4,198 participants. The diseases are grouped into the categories of cancer and metabolic. c., coding sequence change; p., protein sequence change; clin sig, clinical significance according to ClinVar; LP, l ikely pathogenic; P/LP, pathogenic/likely pathogenic; actionable, variant categories considered to be actionable; LOF, loss of function; consequence, the molecular conse quence of the variant; star, ClinVar star status; 2 var, 2 variants must be present, so either homozygous or compound heterozygous for the P/LP variant; homs only indica tes that only homozygotes for the named variant should be reported.

The level of support for each review status in ClinVar decreases from four star downwards (https://www.ncbi.nlm.nih.gov/clinvar/docs/review_status). The great majority of the actionable variants we found were 3 (reviewed by expert panel) or 2 (criteria provided, multiple submitters, no conflicts) star (Table S1). Three of the non-singleton autosomal dominant actionable variants (Table 1) had a review status in ClinVar of 1 star, all of which had a single submitter. The paucity of submissions often indicates rarity of the variant, which in turn makes assessment of pathogenicity and hence actionability more challenging.

### Actionable genotype co-occurrences

We identified four individuals with actionable genotypes in two separate genes (Table 3). We expect some such co-occurrences because of independent assortment, and we should see these according to the product of the allele frequencies of the variants in question. If one or both have drifted upwards in frequency, more will be seen. We found that two of these four Viking Genes individuals had AVs in two different cancer susceptibility genes, while two had actionable genotypes in a cancer susceptibility gene and a metabolic gene.

**Table 3.** Actionable genotype co-occurrences. AD, autosomal dominant; het, heterozygous; hom, homozygous.

| Gene 1 | variant ID | inheritance | genotype | Gene 2 | variant ID | inheritance | genotype |
| --- | --- | --- | --- | --- | --- | --- | --- |
| *BRCA1* | c.5207T>C | AD | het | *PMS2* | c.137G>T | AD | het |
| *BRCA2* | c.6275_6276del | AD | het | *MSH6* | c.742C>T | AD | het |
| *BRCA2* | c.517-2A>G | AD | het | *GAA* | c.841C>T | AR | hom |
| *BRCA2* | c.5073dupA | AD | het | *HFE* | c.845G>A | AR | hom |

### Drifted actionable variants in Northern Isles populations

The isolated Northern Isles populations carry different sets of rare variants compared to cosmopolitan populations. In total, 10 actionable variants across 7 genes (*BRCA1* [MIM: 113705]*, BRCA2* [MIM:600185]*, ATP7B* [MIM: 606882]*, TTN* [MIM: 188840]*, KCNH2* [MIM: 152427]*, MUTYH* [MIM: 604933]*, GAA* [MIM:

606800]) have risen in frequency by at least 50-fold through the action of genetic drift in the Northern Isles (Table 4). These include results published previously on a *BRCA1* breast and ovarian cancer predisposition variant that is 470-fold more common in Orkney than in the UKB^11^, a *KCNH2* Long QT Syndrome variant that is ∼90-fold more common in Shetland^10^, and a pathogenic *BRCA2* variant ∼155-fold more common in Shetland^12^.

**Table 4.** Founder effects on ten actionable variants in Orkney and Shetland. Drifted variants were defined as actionable variants with an increase in frequency over 50-fold higher than UKB, a pedigree origin before c.1800 CE and a minor allele count of ≥5. For UKB, we used the allele frequency browser (based on ∼490k WGS). c., complementary DNA position and nucleotide change; p., protein position and amino-acid change; inher, inheritance; maf.ork, minor allele frequency in Orkney; ork/ukb, fold uplift in frequency in Orkney over UKB; maf.shet, minor allele frequency in Shetland; shet/ukb, fold uplift in frequency i n Shetland over UKB; group, island group the variant has drifted upwards in; origin, subpopulation (parish, isle or region) where the variant arose (by tracing back pedigrees) or has become particularly common.

| gene | c. | p. | disease | inher | maf.ork | ork/ukb | maf.shet | shet/ukb | group | origin |
| --- | --- | --- | --- | --- | --- | --- | --- | --- | --- | --- |
| *BRCA1* | c.5207T>C | p.Val1736Ala | Hereditary breast ovarian cancer | AD | 0.00478 | 470 | 0 | 0 | Orkney | Westray |
| *BRCA2* | c.517-2A>G | . |  | AD | 0 | 0 | 0.00237 | 155 | Shetland | Whalsay |
| *ATP7B* | c.3083_3085delinsG | p.Lys1028fs | Wilson disease | AR | 0 | 0 | 0.00475 | 180 | Shetland | Burra |
| *ATP7B* | c.3007G>A | p.Ala1003Thr |  | AR | 0 | 0 | 0.00356 | 350 | Shetland | Sandsting/Lunnasting/Delting |
| *ATP7B* | c.1772G>A | p.Gly591Asp |  | AR | 0.00215 | 66 | 0 | 0 | Orkney | Westray |
| *TTN* | c.93396_93400del | p.Ala31133_Trp31134insTer | Dilated cardiomyopathy | AD | 0 | 0 | 0.00380 | 3700 | Shetland | Yell |
| *TTN* | c.67519C>T | p.Gln22507Ter |  | AD | 0.00048 | 700 | 0.00071 | 470 | Orkney | West Mainland |
| *KCNH2* | c.1750G>A | p.Gly584Ser | Long QT syndrome | AD | 0 | 0 | 0.00119 | 90 | Shetland | Aithsting, Unst |
| *MUTYH* | c.1205C>T | p.Pro377Leu | *MYH*-associated polyposis | AR | 0.00431 | 70 | 0.00047 | 8 | Orkney | Westray |
| *GAA* | c.1210G>A | p.Asp404Asn | Pompe disease | AR | 0.00287 | 74 | 0 | 0 | Orkney | St. Andrews |

Most actionable variants in the ACMG list have a dominant mode of inheritance and so are actionable if heterozygous, but some are only considered actionable if present in two copies. Table 4 includes five drifted variants in Northern Isles populations that act recessively (also listed in Table 2). These are in *ATP7B* (three drifted variants segregating in our cohorts), *MUTYH* and *GAA*. Wilson Disease [MIM: 277900] is a disorder of copper metabolism, caused by variants in the copper transporting ATPase gene

*ATP7B*. The marked discrepancy between the “genetic prevalence” in populations and the number of clinically diagnosed cases of Wilson Disease is probably due both to reduced penetrance of many *ATP7B* mutations, and failure to diagnose patients with this highly treatable disorder^19^. We identified three founder effects on recessive *ATP7B* Wilson disease variants, two in Shetland, which gives rise to substantial risk of homozygosity or compound heterozygosity. The combined allele frequency in Shetlanders is 0.8%. Together with five other alleles segregating at lower frequencies, the sum Pathogenic/Likely Pathogenic (P/LP) allele frequency is 1.12%; thus about 1/45 Shetlanders are carriers of P/LP *ATP7B* variants. Under random mating, this predicts about 1/8000 will be homozygotes or compound heterozygotes, at risk of being affected. Indeed, one homozygote for the drifted variant c.3007G>A was found.

There is a founder effect on a recessive *MUTYH* variant (c.1130C>T) in Orkney (0.86% carriers, 70x more common than UKB), causing *MYH*-associated polyposis [MIM: 608456] (Table 4). While not increased in frequency compared to other populations, there are two further *MUTYH* variants segregating in the Orcadian population, bringing the combined carrier frequency to 2.9%. This suggests that 1/35 Orcadians are carriers and, assuming random mating among Orcadians, an expectation that ∼1/5000 will be at risk of *MYH*-associated polyposis. Consistent with this, we did observe a compound heterozygote in the Orkney data.

Finally, there are also founder effects in Shetland on two dominant loss-of-function titin (*TTN*) variants, increasing the risk of dilated cardiomyopathy [MIM: 604145] (Table 4). One of these, c.93396_93400del, is >3,000-fold more common in Shetland than in UKB, where there is only a single instance in 917,682 alleles. Together with a third returnable *TTN* variant that does not meet our thresholds to be defined as drifted, the combined allele frequency is 0.5%, meaning 1/100 Shetlanders are carriers. In total, we observed 24 carriers of *TTN* loss-of-function (LoF) variants (23 of which are accounted for by these three alleles).

### An ultra-rare drifted titin variant

The penetrance of the *TTN* alleles included in ClinVar as pathogenic or likely pathogenic is variable and age related. However, the deletion of 5 nucleotides c.93396_93400del causes a frameshift, which creates a premature translational stop signal (p.Trp31134*), predicted to result in an absent or disrupted protein product. This variant is in the A-band of the *TTN* gene. Truncating variants in the A-band of *TTN* are significantly overrepresented in patients with dilated cardiomyopathy (DCM), and are considered to be likely pathogenic for the disease^20^. TTN truncating variants (TTNtv) located in exons highly expressed in the heart (proportion spliced in [PSI]>0.9, or hiPSI), such as this one, increase the odds of DCM by 11-19-fold^21^. This variant has therefore been classified as Likely Pathogenic in ClinVar. We examined data from 347 individuals ascertained with DCM and found to carry hiPSI TTNtv in DCM cohort studies from Europe, USA and Australia, and also interrogated data from TTNtv carriers in the UKB^22^, the Geisinger MyCode Biobank and the PennMedicine Biobank (168 affected with DCM, 2,409 unaffected)^21^. In total we observed 1,200 distinct hiPSI TTNtv in 2,094 individuals. The c.93396_93400del variant was observed twice (1 in UKB and 1 in MyCode), both times in individuals without a diagnosis of DCM. The variant was not observed in the 100,000 Genomes Project^23^ (670 DCM cases, 62,330 non-DCM participants) or All of Us^24^ (245,460 total participants, of whom 125,860 were reported as European heritage), and therefore can be considered ultra-rare.

Analysis of linked data from the EHR for cardiomyopathy (ICD-10 code I42), atrial fibrillation (ICD-10 I48) or heart failure (ICD-10 I50) shows that the 16 carriers of the drifted *TTN* variant c.93396_93400del we identified in VIKING I are nearly four times more likely (P=0.02, Fisher’s exact test) to have an inpatient or day patient record for cardiomyopathy (1/16), atrial fibrillation (2/16) or heart failure (2/16) than non-carriers (4/16 carriers have entries for at least one of these conditions, i.e. a nominal combined penetrance of 25%, versus a prevalence of 6.8% in 4,133 individuals without *TTN* LoF variants). Analysis of ECG data collected in the VIKING 1 recruitment clinic from 15 of the carriers also shows an over-representation of features such as first degree atrioventricular block (3/15) or right bundle branch block (4/15), affecting a total of 7 carriers. In one Shetland *TTN* c.93396_93400del family investigated clinically, the condition has typically been detectable on echocardiography around age 30, but symptoms have not developed until later. One patient required a cardiac resynchronisation therapy-defibrillator, while another is asymptomatic despite an abnormal echocardiogram in their late 50s. Together with the 7 entries in ClinVar submitted to date, we now report this further direct evidence for pathogenicity of this ultra-rare drifted *TTN* variant.

### Impact of sub-population gene pools

The drifted variants are not shared between the two archipelagos, except *TTN* p.Gln22507Ter, which is observed in Shetland among individuals with recent Orcadian ancestry; and *MUTYH* p.Pro377Leu, for which the Shetland carriers have no recent Orcadian ancestry. This latter variant may have been introduced to the Northern Isles more than once, or there may be unrecorded non-parental events connecting the Orcadian and Shetlandic kindreds.

The great majority of these actionable variants can be traced back to individual isles within Orkney or Shetland, or in some cases individual parishes: 13-15 ancient geographic, legal, and religious subdivisions of the Mainlands of Orkney and Shetland (Figure 1). Among published variants, 95% of carriers of the drifted *BRCA1* variant traced their genealogies to Westray, in the North Isles of Orkney^11^, while 78% of the drifted *BRCA2* variant carriers trace back to Whalsay, an isle in Shetland^12^. Likewise, drifted *KCNH2* variant carriers from Viking Genes and NHS Clinical Genetics^10^ trace back either to the parish of Aithsting in the West Mainland of Shetland or Unst in the North Isles of Shetland (Figure 1 and Table 4).

**Figure 1.**
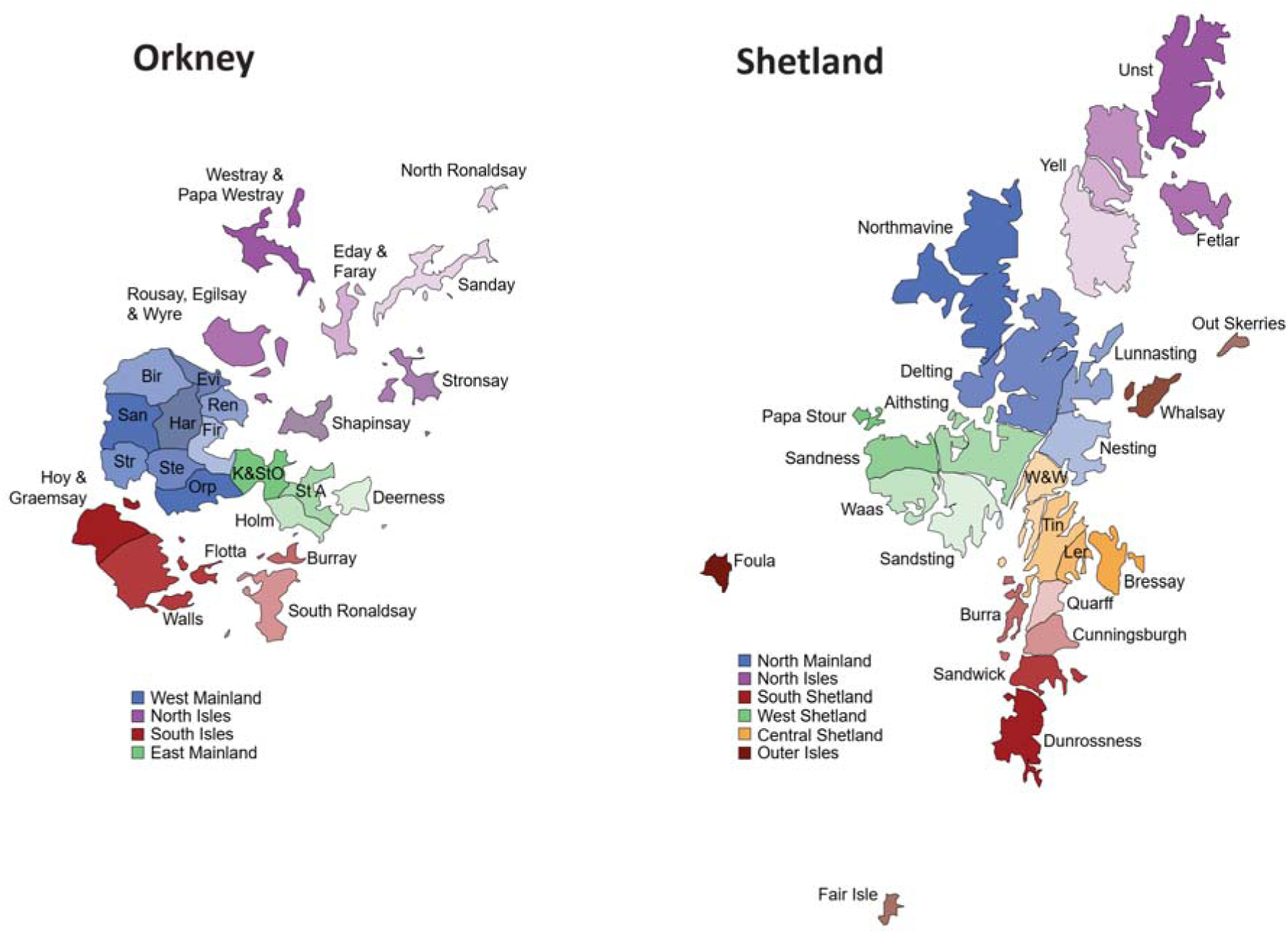
Isles and parishes of Orkney and Shetland. Each archipelago consists of a larger island called the Mainland, with surrounding smaller isles. Both island groups can be divided into 4-6 larger areas, such as West Shetland (known locally as the West side) or the West Mainland of Orkney, which in turn are organised into 25 parishes or isles. Founder effects are usually concentrated in one parish/isle or in one of the larger contiguous areas. W&W, Whiteness & Weisdale; Tin, Tingwall; Ler, Lerwick; St A, St. Andrews; K&StO, Kirkwall & St. Ola; Orp, Orphir; Ste, Stenness; Str, Stromness; San, Sandwick (Orkney); Bir, Birsay; Evi, Evie; Ren, Rendall; Fir, Firth; Har, Harray.

Among the drifted variants reported here, 80% of the carriers of the most drifted *ATP7B* variant have genealogies going back to Burra Isle, Shetland and there is also a strong founder effect on the most highly drifted *TTN* variant c.93396_93400del, this time in Yell, in the North Isles of Shetland, with 100% of carriers tracing their family trees back to there. Further fine scale, within-archipelago population structuring of clinically important variants is observed in Westray, in the North Isles of Orkney, with another *ATP7B* variant having drifted to high frequencies there, and 100% of carriers tracing pedigrees there, similarly to a *MUTYH* variant, where 60% of carriers have Westray genealogies; indeed, we observed a compound heterozygote with ancestry from that isle.

### Genetic down-drift

It is more difficult to quantify which variants have drifted down in frequency, but the near absence of familial hypercholesterolaemia (FH) [MIM: 143890 and MIM:144010] variants is notable, with only one carrier across >4,000 subjects. This *LDLR* [MIM: 606945] variant was observed in Shetland and is the known p.C163Y West of Scotland founder variant^25^; it may have been brought to the isles by recent gene flow from Mainland Scotland; no FH variants were observed in Orkney. No P/LP *APOB* [MIM:107730] or*PCSK9* [MIM: 607786] variants were observed. Significantly fewer volunteers from the Northern Isles carried FH variants than observed in European ancestry participants from the UKB (P<0.0001, Fisher’s exact test). The carrier frequency in the Northern Isles (1/4198) is more than 10-fold lower than that in UKB European ancestry subjects (1/288)^26^.

### The Return of Results (RoR) process

With reference to the Association of Clinical Genomic Science (ACGS) guidelines (https://www.acgs.uk.com/media/12533/uk-practice-guidelines-for-variant-classification-v12-2024.pdf) and local clinical knowledge, four variants in a total of 10 genotype carriers were not deemed returnable by the NHS clinical team (Table 5 and Table S1). Our next goal was to implement a process of return of results (RoR) to carrier participants, summarised in Figure 2. Key principles include: the use of informed consent; respecting the right not to know; checking the research results as far as possible before sharing a potentially actionable variant ID; decisions on whether to return a variant result taken by NHS clinical experts; counselling provided through the NHS, and verification of all the research results in an accredited clinical laboratory. The consent form and RoR participant information sheets sent to cohort members are provided as supplemental information. Frequently asked questions about return of results are available for participants on the study website at https://viking.ed.ac.uk/for-viking-genes-volunteers/faqs/return-of-results

**Figure 2.**
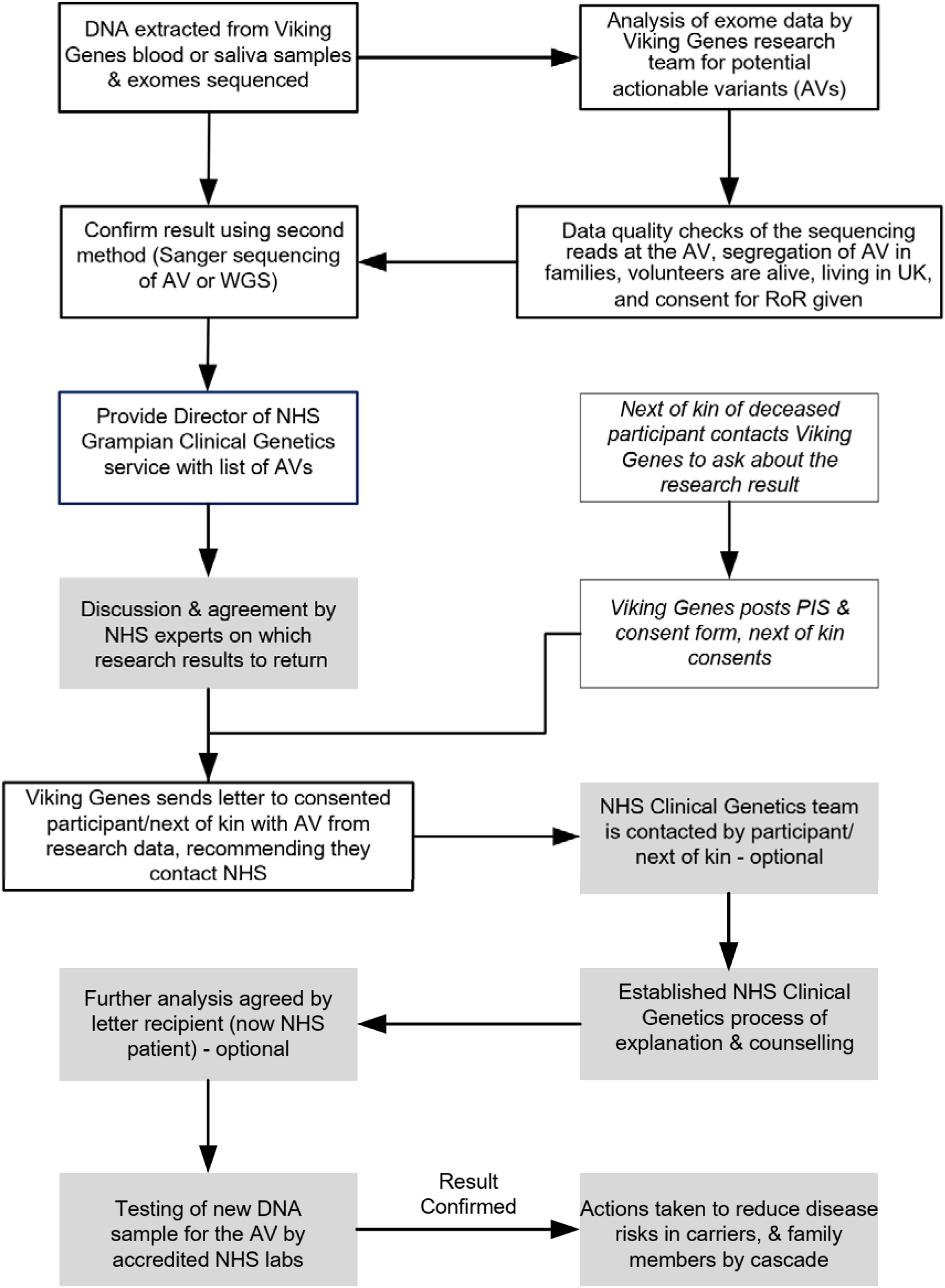
Overview of Viking Genes return of results. Processes implemented by the Viking Genes research team are boxed with no shading; processes implemented by the NHS clinical team are shaded. PIS, participant information sheet (see Supplemental information).

**Table 5.**
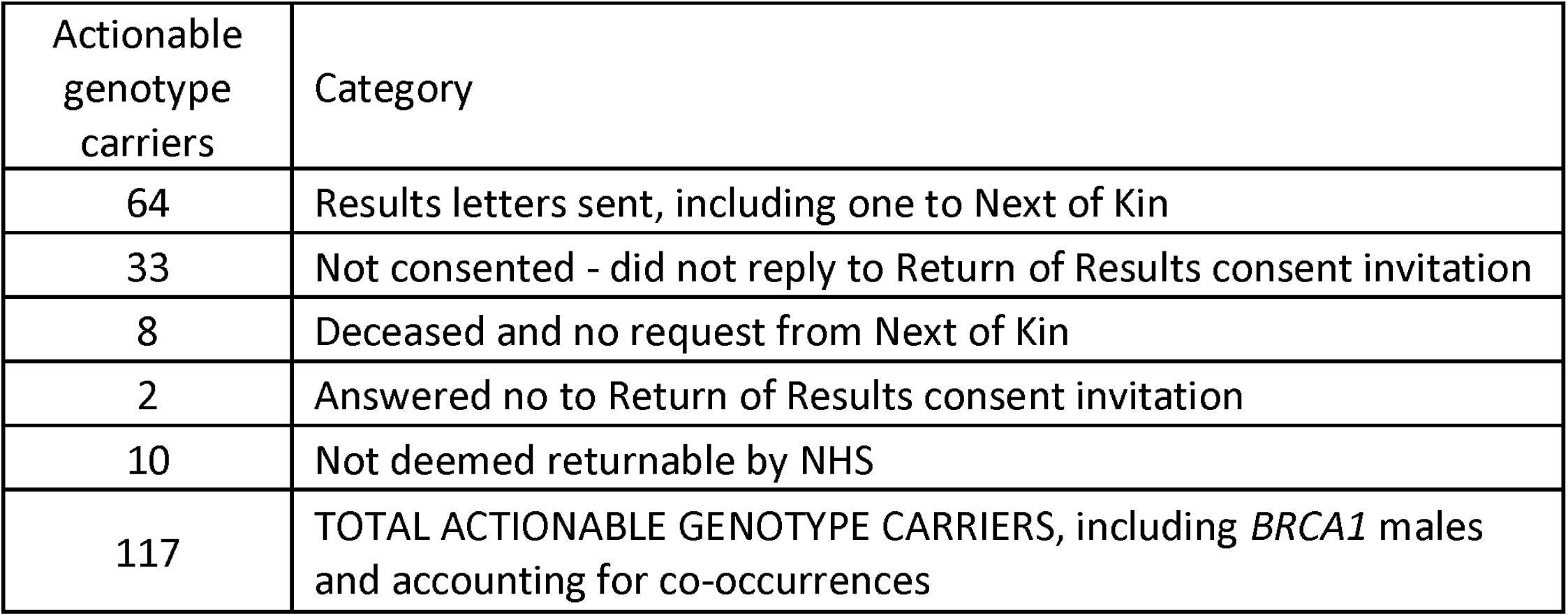
Summary of carriers of actionable genotypes and results returned. Consent was only sought 8-18 years after recruitment, which is probably why 28% of actionable genotype carriers did not reply to the invitation. A further 11% were deceased, while 2.7% replied choosing not to have return of results. NHS, National Health Service.

| Actionable genotype carriers | Category |
| --- | --- |
| 64 | Results letters sent, including one to Next of Kin |
| 33 | Not consented - did not reply to Return of Results consent invitation |
| 8 | Deceased and no request from Next of Kin |
| 2 | Answered no to Return of Results consent invitation |
| 10 | Not deemed returnable by NHS |
| 117 | TOTAL ACTIONABLE GENOTYPE CARRIERS, including *BRCA1* males and accounting for co-occurrences |

We obtained a favourable opinion from the Research Ethics Committee to return results of genotypes that have the potential to improve the health of the individuals carrying them. We subsequently obtained further permission to contact male carriers of pathogenic *BRCA1* variants. This was justified by the fact that although there is no clinical action to be taken with respect to the letter recipient, the information could be of value to their female relatives, who may not be Viking Genes research participants. There are 13 male carriers of the *BRCA1* p.V1736A variant in the ORCADES cohort^11^ and none in VIKING I, so addition of these participants increases the total number of actionable genotypes from 108 to 121. Since there are four individuals with two actionable genotypes each (Table 3), this corresponds to a total of 117 total actionable genotype carriers (Table 5).

We further obtained ethical permission to offer actionable results to the next of kin of deceased participants, if we found any that might have been relevant to the health of the participant who died, and therefore potentially their family, during our research (Figure 2). To date, six next of kin have provided their consent to receive this information, with one actionable genotype disclosed.

The process we have implemented is in good accordance with a checklist of components for return of results developed by an interdisciplinary panel of European experts^27^. In Viking Genes, the return of medically actionable genetic results to participants is optional, i.e. an opt-in approach. This has legal, philosophical, and ethical foundations, to allow patient autonomy and the right not to know. This is achieved at three separate points in the process of implementation (Figure 2). Letters containing results are only sent with the consent of the recipient. Upon receipt, the participant can decline any further investigation by not contacting the NHS about the research finding. Finally, after genetic counselling, the patient can decide not to have the research result confirmed in a clinical setting. It should also be noted that a participant can get in touch with Viking Genes at any time to request to change the consent they gave about RoR, in either direction, or to withdraw from the study. The research team communicates with Viking Genes volunteers by regular newsletters and social media, and invites them to complete occasional research surveys. A subset of participants in the study forms the public - participant involvement (PPI) group. We therefore aim to ensure both that understanding of the opportunity to receive results is high, and the processes we are implementing have the support of the study participants and the populations they are drawn from.

### Initial outcomes from issuing RoR letters

The text of the letters alerting the volunteer to the research result is written in lay language, agreed by the research and NHS teams, and the template document was given a favourable opinion by the Research Ethics Committee. The NHS clinic team is not made aware of the identity of the research participants, but approves return of the variants, which are also noted in the letters. Letters were not sent to all carrier participants at one time. Instead, the 64 RoR letters issued (Table 5) were posted over the course of almost exactly one year. This was designed to prevent the NHS clinics from being overwhelmed, but has meant that outcomes are still being accumulated. For example, it is too early to tally the number of family members contacted by cascade testing from each index case. In the NHS, each index case on average leads to three subsequent cascade tests, half of which are positive^28^. In the case of the Shetland *BRCA2* founder variant c.517-2A>G, a total of 56 people were eligible for cascade letters from just 5 consented carriers in Viking Genes, suggesting a higher level of knowledge of family structure among our participants. We observe that RoR letter recipients take a variable length of time to decide whether to contact the NHS, and not all choose to do so.

In all three of the previously reported actionable founder variants^10–12^, we identified carriers in our research populations who could not have been ascertained from oral history-based cascade testing. Likewise, many of the index cases described here were found to be in family groups not previously recorded by the clinical geneticists. Out-patient NHS EHR data showed that only 9% of our carriers had interacted with clinical genetics, compared with 3.6% of the non-carriers, a 2.5-fold increase (P<0.002, χ^2^). This provides further evidence that a substantial number of the research participants who received a return of results letter were previously unaware of the genomic medicine finding, and have the opportunity for preventative medical intervention as a result of the Viking Genes return of results programme.

## Discussion

### Survey of Actionable Variants

We observed actionable genotypes for 39 autosomal actionable variants, 34 dominant and 5 recessive, and no X-linked variants. Ten founder variants showed strong enrichment, having risen >50-fold in frequency in Orkney or Shetland, while otherwise commonly seen variants, notably those causing familial hypercholesterolaemia, were at least ten-fold lower in frequency. Importantly, both *TTN* and *ATP7B* contain multiple drifted variants, which sum together to even higher frequencies of (likely) pathogenic variation.

The ten actionable variants which have drifted up to much higher frequencies in Orkney or Shetland than in the general UK population mostly have a clear origin in a particular parish or isle within the Northern Isles, 200 or more years ago, highlighting the continuing clinical importance of these historic sub-population gene pools. As is the case with other genetic isolate populations^29^, the combination of multiple founder variants can result in a significant summed population frequency, sometimes in conjunction with non-drifted variants. At the same time, other variants have drifted towards loss, balancing the genetic burden of disease risk.

### Comparisons with other populations surveyed for actionable variants

Overall, these two UK isolate populations have a similar percentage of actionable genotypes (2.5%), compared to cosmopolitan populations worldwide. However, the number carrying one or more actionable genotypes depends on the gene list, rules applied (including bespoke curation) and date of analysis. The ACMG list of recommendations of genes and phenotypes is updated annually, whereas ClinVar is designed to enable the ongoing evolution and development of knowledge regarding variations and associated phenotypes, and its website is updated weekly. The status of a variant, particularly if observed rarely worldwide, may therefore change over time as more data are submitted, interpreted and reported in ClinVar. This makes it challenging to compare our Viking Genes actionable variant carrier percentage with populations reported in other publications. In the first 50,000 exome sequences from the UKB population cohort, an actionable genotype figure of 2.0% was reported^2^. However, that analysis used v2.0 of the ACMG list^30^, containing 59 genes, rather than the 81 in v3.2 that were surveyed here. Similarly, a figure of 2.8% can be calculated from the DiscovEHR study^31^ and 2.3% in Iceland^32^, using the ACMG list v2.0. Another way to compare with studies in other populations is to use the ACMG 2.0 (59 gene) list to assess our data in the Northern Isles population, in which case the percentage with actionable genotypes drops to 1.4%.

The inclusion of *HFE* [MIM: 613609] p.Cys282Tyr homozygotes, in later versions of the list, would on its own increase the percentage of actionable genotype carriers in the UKB from 2.0% to 2.6%. This is based on the *HFE* variant having a reported homozygous frequency of 0.6%, or 1 in 156, in the UKB^33^. We therefore show here that the Northern Isles populations have an AV percentage that is in a similar range to the rest of the population of the UK as sampled in the UKB, to the DiscovEHR study of people in Pennsylvania, USA, and to the Icelandic population (4.0% in the most recent analysis, including manual curation)^32^, but much lower than that reported for the Old Order Amish of Pennsylvania^34^.

We identified a sufficient number of carriers to provide specific evidence for pathogenicity of one drifted variant in *TTN*, c.93396_93400del, p.Trp31134*. A total of 44 genes were recently asserted to be implicated in non-syndromic Dilated Cardiomyopathy (DCM) by the ClinGen DCM Gene Curation Expert Panel^35^. *TTN* is one of 11 genes with definite evidence, and featured in a gene-based analysis of penetrance and clinical phenotype in 18,665 UKB participants^36^. A sub-analysis for genes including *TTN* revealed DCM or early DCM features in 45.4% of cases^36^. Moreover, significant excess mortality was observed among carriers of *TTN* truncating variants in Dutch founder pedigrees, driven by subjects ≥60 years^37^.

### Return of Results to Research Volunteers - Benefits and Challenges

The return of results processes that we successfully implemented for Viking Genes participants as described here were somewhat more straightforward than they would be in cosmopolitan populations, due to the reduced number of different actionable variants in the ancestral populations of the Northern Isles of Scotland, and the high support for and trust in scientific research in these communities. Nonetheless, the processes and experiences of returning actionable genetic results in Viking Genes are directly relevant as an exemplar (especially in the UK) for other research cohorts interested in the feasibility and logistics of the return of actionable results to their participants. Our recent recruitment to a new cohort study, VIKING II, was eligible to people of Northern Isles ancestry regardless of domicile^15^. At the outset of recruitment, VIKING II offered the option of consent to return of selected clinically actionable results. This option was chosen by an overwhelming majority (98%), 6,054 of the 6,178 participants who consented and completed the study questionnaire. This is in good agreement with several other studies that also show strong support for receiving actionable findings, both in theory and in practice^38,39^. These views may mean that the return of actionable genetic findings from research will in future become the default practice^39^. However, the costs, harms and benefits of returning actionable results are still at an early stage of being fully characterized.

For some diseases in the ACMG list, there can often be a long delay before accurate diagnosis, due to the overlap of symptoms with other more prevalent conditions. Examples we have found in Viking Genes include Pompe disease [MIM: 232300], caused by a deficiency of the enzyme acid alpha-glucosidase (GAA), which can lead to nerve and muscle problems, Wilson disease [MIM: 277900], caused by copper overload, and the iron overload condition hereditary haemochromatosis [MIM: 235200] (*HFE*). Early diagnosis and thus early intervention can have an impact on future quality of life. Furthermore, in addition to the medical intervention available for each actionable result, a clearer diagnosis/etiology following the return of results can also be of value, for those already affected, particularly for conditions like cardiomyopathy which have complex symptoms and presentation. Increased awareness of these otherwise rare conditions among healthcare workers in the Northern Isles could lead to decreased times to diagnosis, with concomitant benefits to patients.

Cross-sectional approaches, for example to understand the penetrance of rare-variants in cardiomyopathy-associated genes, can provide valuable data^40^, and genome-first evaluation at scale can enable new diagnoses missing from clinical healthcare^41^. However, the scientific literature is only just beginning to provide insight into how the ACMG guidelines have been translated into precision health outcomes for the recipients of the findings: for some genes benefits clearly outweigh harms, while for others there is little evidence as yet^42–44^. This is likely to gather pace in the near future, as more large scale initiatives such as All of Us^3^, the Healthy Oregon Project^45^ and the Colorado Center for Personalized Medicine^46^ implement the return of actionable results. Furthermore, disease manifestation, healthcare outcomes and costs of disclosure have recently been described for actionable findings in a UK setting from the Genomics England 100,000 Genomes Project^47^. Genomics England returned results for actionable genotypes in 13 genes within the 81 genes on the ACMG3.2 list, for hereditary cancer syndromes and familial hypercholesterolaemia, plus carrier status for cystic fibrosis ([MIM: 219700], not on the ACMG list). Due to a lack of consensus on benefit versus harm and the challenges of reduced penetrance, Genomics England chose not to inform carriers of actionable genotypes increasing the risk of Long QT syndrome, cardiomyopathies, aortopathies, further inherited cancers, metabolic diseases and the most frequent genetic disease, hereditary haemochromatosis (www.genomicsengland.co.uk/initiatives/100000-genomes-project/additional-findings).

Jensson *et al*^32^ found shorter median survival among persons carrying actionable genotypes than among non-carriers, in Iceland. Specifically, they report that carrying an actionable genotype in a cancer gene on the ACMG list was associated with life span that was three years shorter than that among noncarriers, with causes of death among carriers attributed primarily to cancer-related conditions. Furthermore, while many studies have demonstrated the diagnostic and therapeutic value of exome or whole genome sequencing in critically ill paediatric cases, a recent study shows that the diagnostic utility of exome sequencing in critically ill adults is similar, largely due to uncovering variants in medically actionable genes^48^.

Our data highlight the clinical relevance of local genetic sub-populations, as most of the founder variants are also associated with one or other parish or isle, with deep genealogies linking most carriers to ancestors from there (notwithstanding ancestral non-parental events which obscure the connection between genetics and geography). Within each archipelago, it is natural that historic barriers to marriage in the form of storm-prone, strongly tidal ocean and sea channels and sounds, have given rise to subpopulation gene pools or micro-isolates^7,49^. Within the main islands, the parishes are sometimes separated by low hills or sea firths, and sometimes not, but marriage records show they were demographically important over the centuries.

### Population Screening Opportunities

Population screening in a clinical context is currently available in some populations that show founder effects, such as an NHS programme offering *BRCA*-testing to those with Jewish ancestry (https://www.nhsjewishbrcaprogramme.org.uk/). Charitable organisations are also increasingly making specific genetic tests available to populations of higher risk. Examples include improving the prevention and diagnosis of Jewish genetic disorders in the UK (https://www.jnetics.org/) and the provision of postal tests for genetic haemochromatosis (https://www.haemochromatosis.org.uk/).

Our estimates of the frequency of the *BRCA1* and *BRCA2* actionable variants in Westray and Whalsay, respectively, (1/19 and 1/43)^12^ show that the local gene pool frequencies can be much higher than the archipelago-wide frequencies. This in turn suggests that individuals with ancestry from islands such as Yell, Burra and Westray are at higher risk of being carriers than the Shetland- or Orkney-wide data suggest. The ability to target testing to those with an origin in a particular parish may improve the cost-effectiveness of future screening. At the same time, with increasing movement, younger generations and inhabitants of the towns of Kirkwall and Lerwick (and of course the diasporas outside the Northern Isles) tend to have more mixed ancestry, so the importance of these micro-isolates is waning.

We report four actionable genotype co-occurrences. Many more instances in other populations are likely to be uncovered in the future, as genetic analyses move from limited gene panels towards WES and WGS. A recent review, for example, describes Multi-locus Inherited Neoplasia Allele Syndrome (MINAS), which refers to individuals with germline pathogenic variants in two or more cancer susceptibility genes^50^.

We have not investigated LoF variants that may be present in our data, but which are not reported in ClinVar. There may be potentially important ultra-rare variants in this class, which warrants future research. Furthermore, clinically relevant variants that lie outside of exomes, and structural variants, may be uncovered in subsequent research. However, except for the most drifted variants, the assessment of pathogenicity will always be limited by the modest numbers of carriers likely to be found in our cohorts. We also note that the high levels of engagement and knowledge of kinship led to higher uptake of cascade testing, and thus a higher-than-expected demand on clinical services, compared to previous cascade testing statistics reported in the NHS^28^.

Studies of population genetic structure using unselected genome-wide markers^7,51^ are not always considered relevant for clinical genetics. However, a number of reports now demonstrate the consequences of such population structure for clinical genetics at the population level. A case in point is the Irish Travellers, shown to be a distinct population isolate of Irish origin^52^ with their own suite of Mendelian conditions at high frequencies^53^. Similarly, the genetic isolation of Orkney and Shetland has been shown to translate into clinically important differences at the population level, from the earliest studies of Northern Isles genetics, showing a founder effect on the *SOD1* [MIM: 147450] p.D90A allele^54^, to novel Mendelian conditions^55–57^, and distinct blood group/isozyme^58^ and uniparental^6^ markers. More recently, numerous founder effects, both on Mendelian recessive alleles^8^, and on dominant actionable variants^10–12^ have been described.

Orcadians and Shetlanders thus belong to a group of European-heritage populations, including Ashkenazi Jews and Irish Travellers, where high frequencies of important clinical variants have arisen from genetic drift, with concomitant simplification of the gene pools providing valuable opportunities for population-wide, cost-effective targeted genetic screening.

### Data and Code Availability Statement

There is neither Research Ethics Committee approval, nor consent from Viking Genes participants, to permit open release of the individual level research data underlying this study. The datasets generated and analysed during the current study are therefore not publicly available. Instead, the research data and/or DNA samples are available from on reasonable request, following approval by the Data Access Committee and in line with the consent given by participants. The ClinVar-exome pipeline code is available on GitHub (https://github.com/viking-genes/clinvar_pipeline). Frequencies for the ten drifted variants reported here were derived from the UK Biobank Whole Genome Sequencing (WGS) project and were obtained from the publicly available UK Biobank Allele Frequency Browser (https://afb.ukbiobank.ac.uk/), which was generated by the WGS consortium under the UK Biobank Resource (project ID 52293).

## Data Availability

The datasets generated and analysed during the current study are available from on reasonable request, following approval by the Data Access Committee and in line with the consent given by participants. The ClinVar-exome pipeline code is available on GitHub (https://github.com/viking-genes/clinvar_pipeline).

## Acknowledgments

This work was funded by the Medical Research Council University Unit award to the MRC Human Genetics Unit, University of Edinburgh, MC_UU_00007/10 and a Wellcome Trust Institutional Translational Partnership Award (University of Edinburgh 222060/Z/20/Z -PIII031). LK was supported by an RCUK Innovation Fellowship from the National Productivity Investment Fund (MR/R026408/1). ORCADES was supported by the Chief Scientist Office of the Scottish Government (CZB/4/276 and CZB/4/710), a Royal Society URF to JFW, and Arthritis Research UK. JSW was supported by the Medical Research Council (UK), Sir Jules Thorn Charitable Trust (21JTA), British Heart Foundation (RE/18/4/34215), and the NIHR Imperial College Biomedical Research Centre. This research has been conducted using the UK Biobank Resource under Application Numbers 47602 and 19655.

Emily Weiss and Reka Nagy assembled the Orkney pedigree, and Barbara Gray the Shetland pedigree, using records at the General Register Office, the Shetland Family History Society, and study information, building on earlier pedigree work by Ruth McQuillan and Jim Wilson. Sanger sequencing was performed by the technical services team at the MRC Human Genetics Unit. We thank the NHS Grampian clinical genetics team and the Viking Genes research team (in particular David Buchanan, Rachel Edwards and Craig Sinclair) for their contributions to the implementation of the return of results process. Finally, we thank the people of the Northern and Western Isles for their involvement in and ongoing support for our research.

## Author Contributions

SMK managed the project and drafted the manuscript. LK created and implemented the ClinVar-exome pipeline, with input and support from KJ, and analysed the exome datasets. MDM updated the pipeline. CD validated exome data by PCR and Sanger sequencing of genomic DNA. MH validated exome data by analysis of WGS data. SMK, ZM and JFW planned the RoR methodology. ZM and JD provided information on the clinical epidemiology of the Northern Isles and clinically reviewed the variants to be reported. JSW, SLZ and PKT interpreted phenotype data in cardiomyopathy variant carriers. GT and ARS conceived and managed the Viking Genes exome sequencing. ZM led the NHS RoR clinical team, which included genetic counsellors EC and LS, who discussed actionable results with participants. JFW is the Chief Investigator of Viking Genes, was awarded funding to implement the work, analysed the data and helped draft the manuscript. All authors provided input and feedback on drafts of the manuscript.

## Ethical Approval

Eligible participants were recruited to Viking Genes, NHS Lothian South East Scotland Research Ethics Committee reference 19/SS/0104. Research participants gave informed consent for research procedures including DNA sequencing (mandatory) and return of actionable results (optional).

For the purpose of open access, the author has applied a Creative Commons Attribution (CC BY) licence to any Author Accepted Manuscript version arising from this submission.

## Supplemental Files

Supplemental Table S1 All actionable variants reported by our ClinVar pipeline.

Supplemental Information – RoR Participant Information Sheet (PIS), RoR Consent Form and RoR PIS for Next of Kin.

